# Aerosol blocking assessment by different types of fabrics for homemade respiratory masks: spectroscopy and imaging study

**DOI:** 10.1101/2020.05.26.20100529

**Authors:** Mauricio Foschini, Adamo Ferreira Gomes Monte, Ana Caroline Moreira Mendes, Renata Scarabucci Janones, Alexandre Maletta, Carla Denari Giuliani, Bruna Aparecida Rodrigues Duarte, Kleber Del-Claro

## Abstract

During the COVID-19 pandemic, there is no agreement, until the current date, about the recommendations of homemade face mask use for the general population, and one of the reasons is a lack of information about their real protective rule on spreading aerosols and viruses. This is a comparative study regarding the relative efficiencies of commercial respiratory masks (medical masks) and homemade fabric masks, which may guide authorities across the globe, following the “Advice on the use of masks in the context of COVID-19”, by the World Health Organization. We described two optical methodologies for charactering respiratory masks. It happens that the aerosol scattering coefficient is linear as a function of its concentration inside the mask chamber. Quantitative optical properties of scattering for a large batch fabrication of masks were demonstrated, making the mask N95 suitable for use as a reference standard.

## INTRODUCTION

Since the outbreak of COVID-19 pandemic, the widespread face mask use recommendations varied among different authorities and countries in the globe. While there is agreement about face mask use for symptomatic people and health-care professionals, controversies exist about extending this recommendation to general public, in community settings^1^. Reasons might be, among others: concerns about rational use of medical supplies, and also lack of information about the real protective rule of aerosol spreading by homemade masks.

While speaking, coughing or sneezing, a person eliminates micro-droplets in the air, which may vary in diameter and, thus, behave differently when falling down to a surface or entering other people’s airway, for instance. It is mostly import to make a distinction between particles smaller than 10 μm in diameter and those larger than that, because of their significant qualitative differences including suspension time, penetration of different regions of the airways and requirements for different personal protection equipment (PPE). “Airborne transmission” is then commonly used to mean transmission by aerosol –size particles of < 10 μm ^2^.

In 2003, a new coronavirus emerged in humans, causing a severe acute respiratory syndrome (SARS). This new virus was called SARS-CoV. There are consistent data to consider SARS-CoV an airbone transmission route disease^3–5^. It is possible that the new coronavirus identified in 2019, and nominated SARS-CoV-2, may also be spread by aerosolized particles. However, this was not confirmed until the current date ^6–9^. Nonetheless, considering that a significant portion of individuals with coronavirus lack symptoms (“asymptomatic”), and in order to help slowing the spread of COVID-19, authorities have been recommending home-made masks for general population. The World Health Organization published an “Advice on the use of masks in the context of COVID-19” on April, 6^th 10^.

Considering the urgent need of scientific data regarding the efficacy of homemade masks, comparative studies testing surgical masks, n95 and fabric masks are recently being published ^11^. Right after Brazilian health authorities recommended the use of home-made masks by the general population, in March 2020, different materials and fabrics were used for that purpose, depending on their availability. Therefore, the present study tested papers, fabrics, and nonwoven fabrics, commonly used in home-made masks in Brazil. So, in this work, we pointed out two methods for the assessment of different types of respiratory masks that will be used to demonstrate two distinct methodologies for measuring aerosol passage through the masks.

## MATERIAL AND METHODS

For this study, the following samples were used: N95-PFF2 mask with EUA/INMETRO certification, surgical masks (SKY™ and Embramac™), confectioner mask, 97 % cotton fabric, 100 % cotton fabric, unwoven fabric, 3M Scotch-Brite™ reusable cloth - Multipurpose from 3M, reusable Scotch-Brite cloth - Maximum absorption of oil and water from 3M™, legging fabric, elastane fabric, paper coffee, paper towel (Snob™), paper towel (Yuri™). The samples with their compositions are described in Table 1.

**Table 1.** Sample used with description.

| **fabric** | **M#** | **description** |
| --- | --- | --- |
| N95/PFF2 | M1 | EUA/INMETRO<br>certification |
| Surgery mask | M2 | Sky™ and Embramac™ |
| Confectioner mask | M3 | Several Brands |
| Cotton 100% | M4 | Tricoline fabric |
| Cotton 97% | M5 | Tricoline fabric |
| TNT | M6 | Gramage 40 |
| Multi-use Wipes | M7 | 3M -Scotch-Brite |
| Legging fabric | M8 | Poliamida 87% e<br>elastane 13% |
| Knitted 88 % cotton | M9 |  |
| Knitted 100 % cotton | M10 | 2-layers |
| Coffee filter | M11 | Melitta™ #103 |

Figure 1 shows the experimental setup used in the execution of the measurements to determine the percentage of aerosol retention by the fabric used as homemade masks. A piezoelectric nebulizer (3W power, 108 kHz frequency) was used to create the aerosol from distilled water. The aerosol was transported through a line attached to a vacuum cleaner. A valve for pressure and flow control were added to the transport line. The pressure measurements were obtained with a BMP180 pressure sensor using the Arduino nano V3 - ATMEGA 328p programmed as interface and a computer. The chamber was made in a 3D printer with 6 flat optical windows and provided a mask holder at the central part which can be easily detached for changing the mask, with 3 windows before the sample 3 windows after the mask holder following the flow of the aerosol.

**Figure 1.**
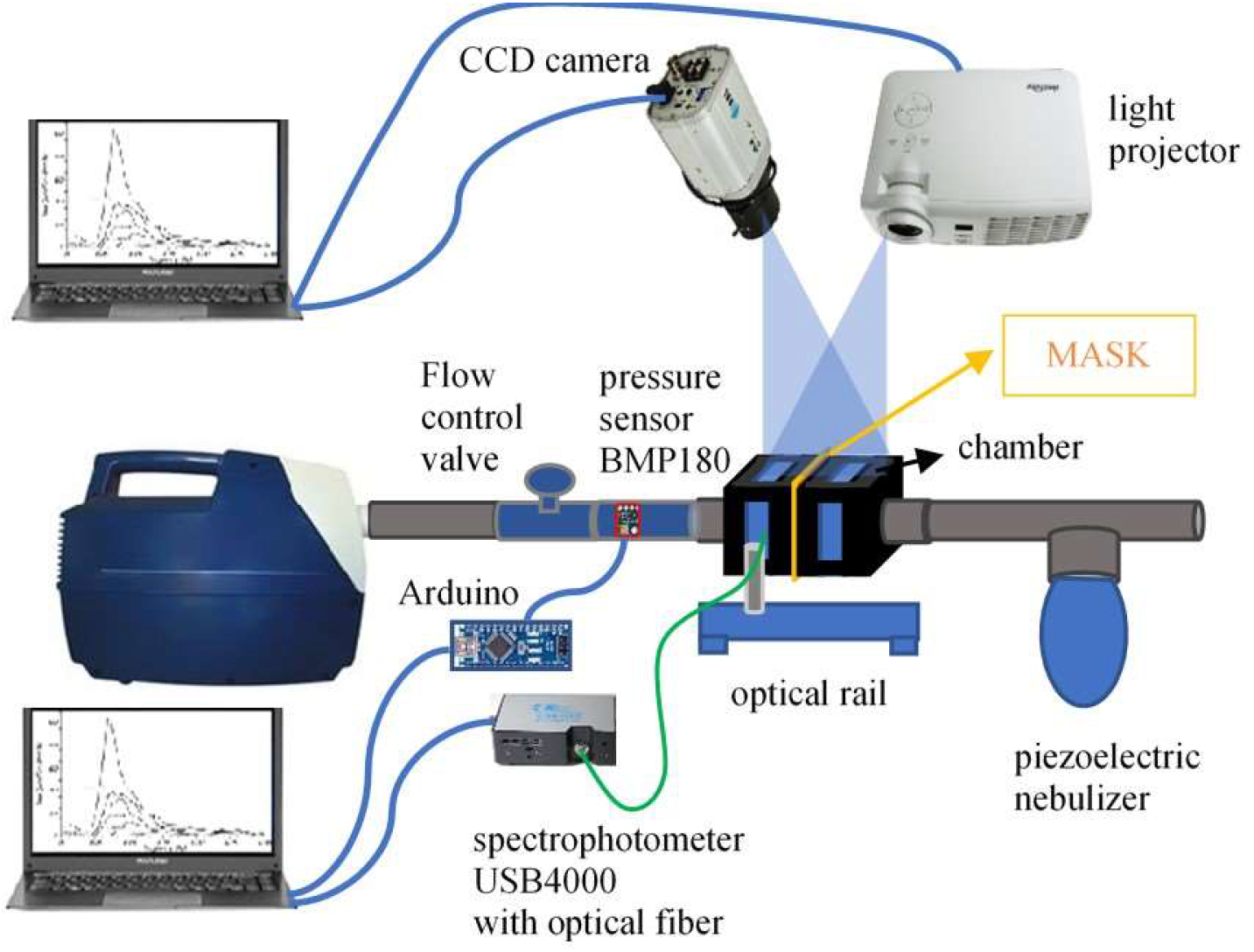
*Experimental setup used to quantify the percentage of aerosol retention on the mask study. The SFDI imaging system is based on a light projector that is used for projecting modulated light images with different spatial frequencies “fx” on a reference sample. Afterwards, a CCD camera collects the image from the diffuse reflectance. After processing the collected images, it is possible to determine the scattering maps (μs’)*.

### Optical and imaging experiments

For the optical scattering measurements two different techniques were provided and used simultaneously, as you can see in Figure 1, comparing the intensity of light dispersion by the droplets between the windows before and after the sample.

The first measurement technique, named technique #1, was performed with the incidence of white light projected on the sample holder at maximum intensity and in the perpendicular windows, the light intensity scattered before and after the sample were measured. The light intensity measurements were performed using the Ocean Optics spectrophotometer - USB4000 and collected with a 600 μm optical fiber, attached to the optical setup that allowed properly alignment for the windows before and after the mask. This measurement was named as technique #1. The second measurement (named technique #2) consisted of measuring the Mie scattering light, obtained by the projection of pattern of images, with certain frequencies, on the mask chamber, and the imaging the capture using a CCD camera. In order to do this, the scattering (μs’) coefficients of the aerosols crossing the masks were carefully measured through the Spatial Frequency Domain Imaging technique (SFDI)^12^.

The SFDI technique employs sinusoidal patterns of spatially modulated light as an excitation source. As shown in Figure 1, the experimental set-up consisted in the compact light source (OSL2, Thorlabs Inc.), a light projector (P300, AAXA Inc.), and one CCD camera 1280 x 1024 (Thorlabs Inc). The light source output is connected through the optical fiber to the DMD (Digital Micromirror Device), which is inside the projector and modulates the light in order to be projected on the mask chamber. The image acquisitions were realized without external light interference as the room was completed dark. These conditions improve signal to noise ratio (SNR) and prevent crosstalk with spurious lights. In order to determine the aerosol scattering properties at a given wavelength, SFDI requires spatial modulation of the illumination as well as the measurement of a well-known optically diffusive phantom used as a reference (reference phantom). The sine wave patterns generated by the DMD module had three different phases (0°, 120°, and 240°) and seven spatial frequencies from 0.04 to 0.28 mm^-1^. These patterns were sequentially projected into the mask chamber and diffuse light was collected with the CCD camera. The original images had 1024 x 1024 pixels in size and it can cover an area of phantom 70 mm x 60 mm.

## RESULTS AND DISCUSSION

Technique #1 obtained with the spectrometer showed that it was possible to measure the spectra by comparing the light intensity scattered by the aerosol just before and after crossing the mask, by using the optical setup as in Figure 1. Considering a TNT mask as an example, the spectral integration in Figure 2-A has provided the relative intensities before and after the masks, and the proportion of those intensities would correspond to the absolute change of particle number inside the aerosol crossing the mask.

**Figure 2.**
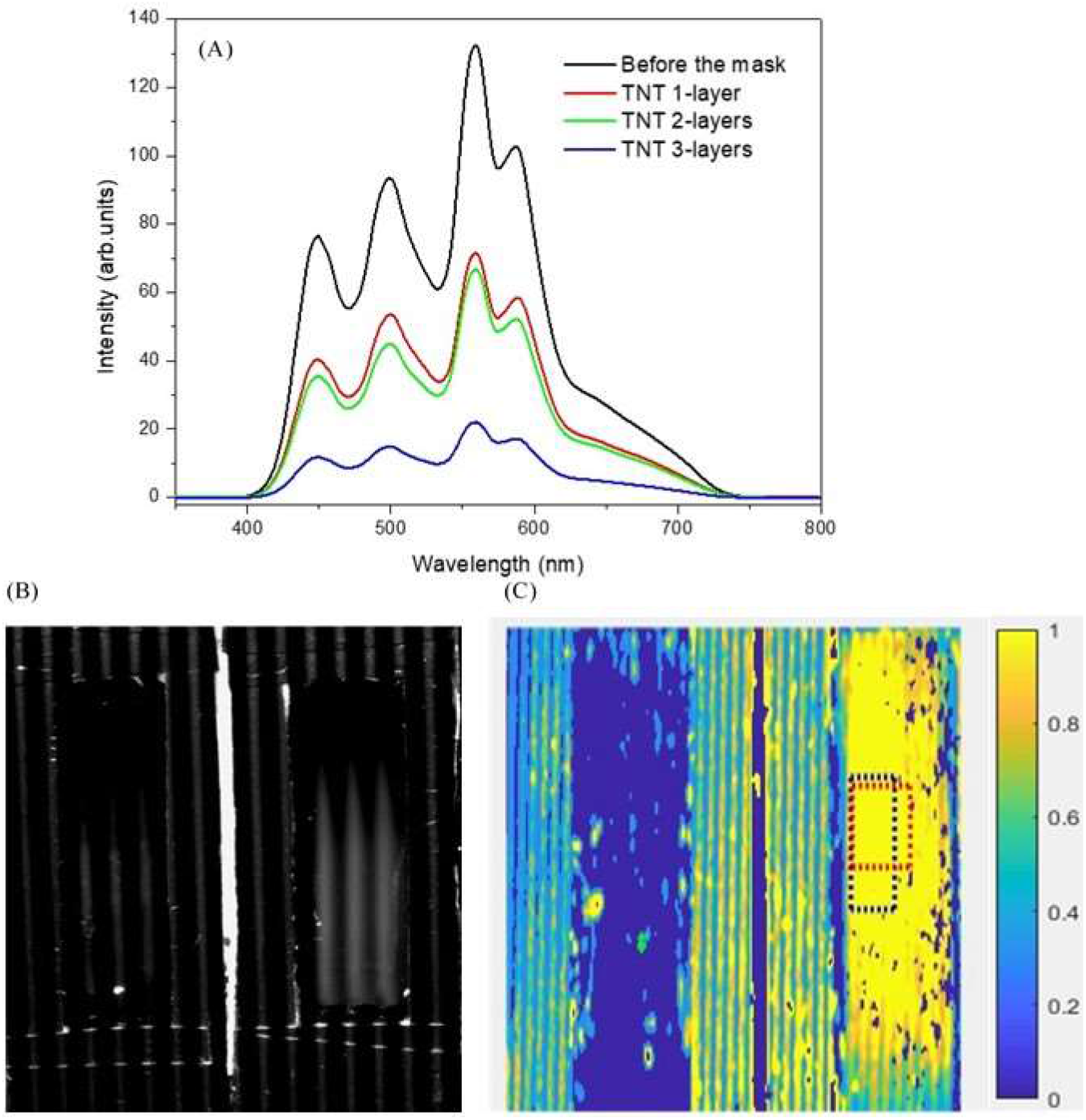
*(A) Optical spectra obtained before the mask and after it, regarding the different layers of TNT mask. (B) The sinusoidal patterns of spatially modulated light on the mask chamber. (C) For comparison reasons a typical spectrum (μs’) was shown for a N95 mask. The yellow color represents the maximum scattering coefficient from aerosol at the right chamber before the mask. The blue color represents the absence of aerosol on the left chamber after the mask*.

In our first set of measurements, the scattering maps spectra were determined from the total diffuse reflection by using the method of SFDI shown early. Figure 2(B) shows that the reduced scattering coefficient (μ_s_’) from the “M” batch masks changed by different cloth types. The following SFDI results are merely computational calculations that might reflect the noise and imperfections related to the recorded images. We understand that the image quality obtained from a sensitive CCD camera with 1024 x 1024 pixels in size is enough to guarantee the accuracy of the calculated u_s_’. As for the SFDI experiments we were able to quantify the optical properties of an unknown mask such as the one in Figure 2(A), based on a known reference phantom with previously calibrated values of μ_a_ and μ_s_’. Thus, the following graphs represented the measured scattering coefficients as a function of the spatial frequency (μ_a_ x *f*_x_) and the six different masks. Note that the interrogated volume was associated to the total number of particles of aerosol.

Thus, Figure 3(A) was obtained with the proportionality of scattered light intensity obtained by spectral integration from light scattered regarding different masks exhibit by Figure 2(A). The first column demonstrated that the n95 mask had the highest aerosol blocking, at least 99.9 %, followed by surgical masks, 99.0 %. However, regarding the efficiency scale, the paper filter also reached good aerosol retention, although it did not worked well providing air passage that also makes the humidity increases. It will be shown that it had the worst performance of pressure test with increasing humidity.

**Figure 3.**
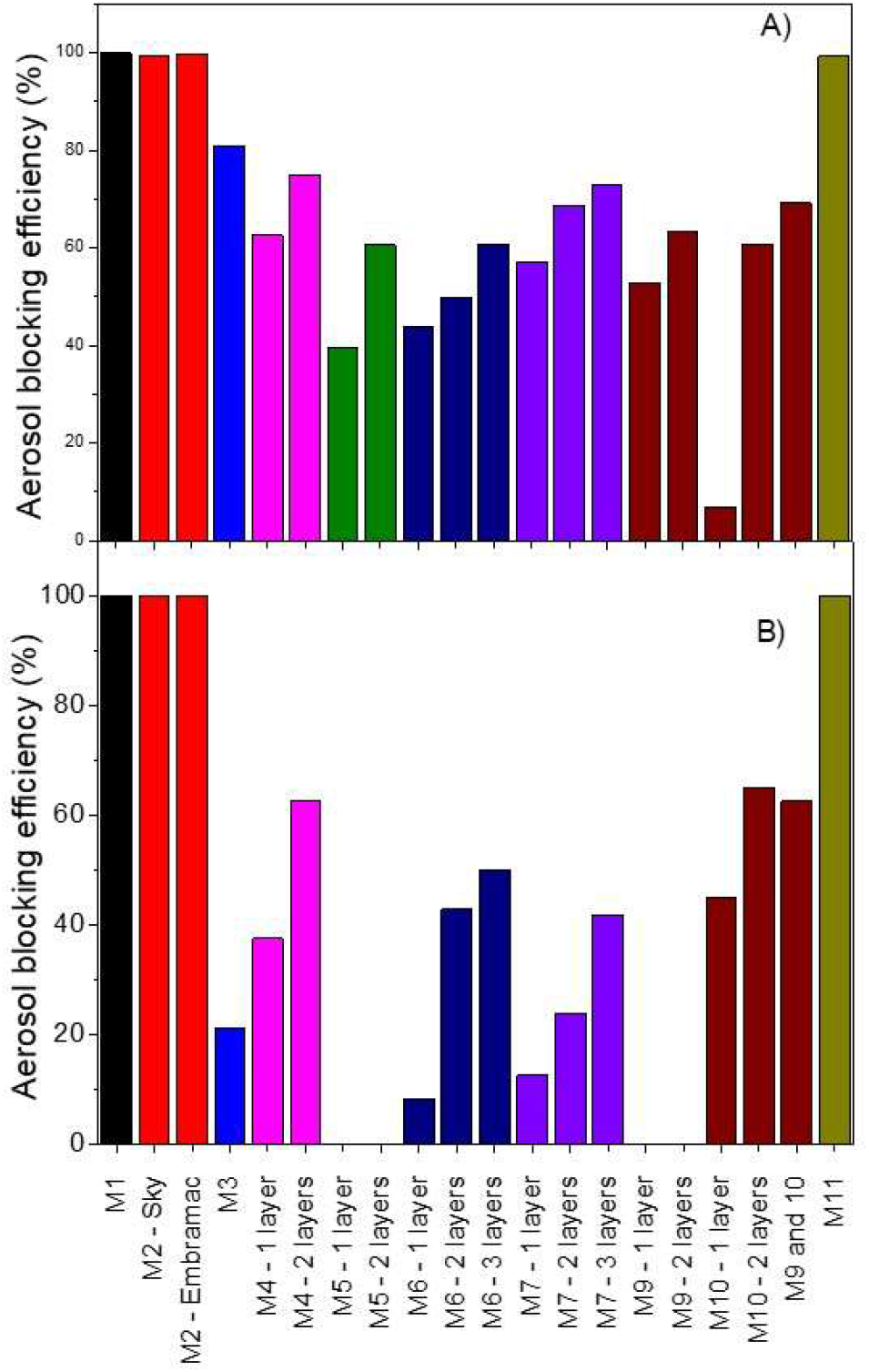
*Aerosol blocking efficiency different type fabrics. (A) scattered light intensity (B) obtained by technique #2. The diffuse reflectance*.

Through total diffuse reflection, using the SFDI method shown previously, in Figure 2 (B), obtained the same behavior in Figure 3(B) obtained by technique #2. The diffuse reflectance components are the inputs to the diffusion equation and are ultimately used to calculate μ_a_ and μ_s_’. By scanning each pixel of the recorded images, entire two-dimensional maps μ_s_’ could be reconstructed. More details of these calculation steps were described in a previous publication^13^. Regarding the analysis, scattering parameters were quantified by fitting an analytical spatial frequency-domain diffuse reflectance model to the measured reflectance, by using a reference phantom with known optical properties to calibrate the instrument prior to every measurement. The projection window that projects the modulated light is focus sensitive so the phantom was maintained in the same focal distance during the measurements, with working distance at 40 cm approximately.

The paper towels performed well, but they had integrity problems during the realization of the measurements, mainly due to the high pressure discontinuity with increased humidity, making its use undisqualifyed. Reusable cleaning cloth towels, commonly used in kitchen cleaning, are similar to TNT, but they have many holes that looks like channels for the aerosol thus decreasing any protection efficiency. Although Figure 3(A) showed a good performance for this type of fabric, since technique #1 used and optical fiber, the measurements are focused on small spots, and this type of fabric might have had large spacings that creates aerosol channels. On the other hand, the technique #2 was more efficient when measuring large regions of the chamber with the masks with larger porosities, since it worked by averaging the image pixels on large areas. In Figure 3(B) it is observed that these fabrics do not perform well, and the image captured during the measurement, as on Figure 4, clearly shows aerosol flow lines and turbulences on the left optical window after the mask. The situation got worse because this fabric was treated to absorb water more efficiently, then the capillaries of the material could bring any solid particles from the outside into the inside, as the humidity increases with the individual’s breathing. Large accumulation of water in the windows after the fabric was observed, as opposed to the surgical masks, since its hydrophobicity created droplets on the incident surface. Finally, any deformation of the fabric associated with usual washing may decrease their durability compared to the 100 % cotton, such as, Tricoline. Therefore, it could become one of the last options of use.

**Figure 4.**
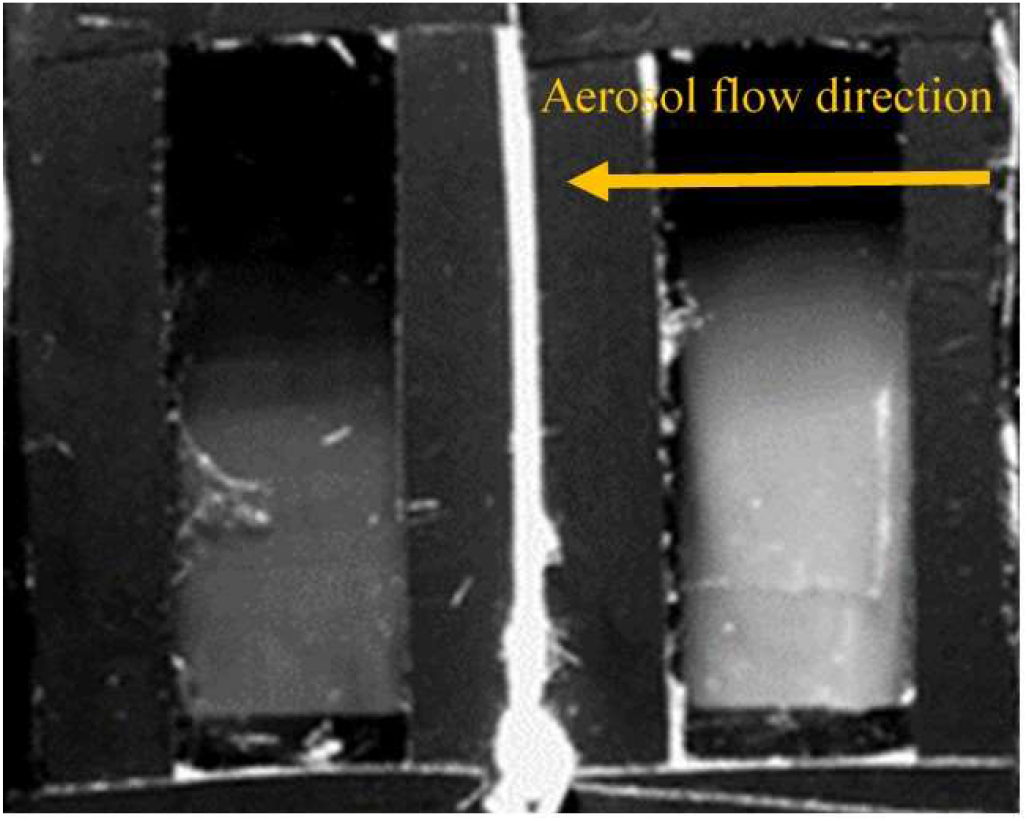
*Clearly shows aerosol flow lines and turbulences on the left optical window after the mask*.

Both techniques showed that fabrics and meshes having some elasticity showed less performance than Tricoline, because the elastic deformations at some point increases the air passage. Cooking masks, despite appearing close to surgical masks and having a treatment to make them hydrophobic, have much less layers than surgical masks, thus reducing dramatically their efficiency, forming aerosol channels as observed in TNT also. The Legging fabric performed well, although it was not added in Figure 3 due to air passage or breathing difficulty. As the aerosol flow increased in the mask chamber, the fabric suffered deformations that could impair its efficiency, making it questionable for the use of protective masks. Afterwards, conventional fabrics with a higher cotton content were more effective, reaching effective values of approximately 70 % as for Tricoline fabric.

Experimental discrepancies sometimes may occur when comparing Figure 3 (A) and (B), regarding each type of mask. However, we were able to provide better results in a sustainable way, e.g., by averaging the results from both techniques, #1 and #2., obtaining the sequence of efficiency of the tissues present in Figure 5

**Figure 5.**
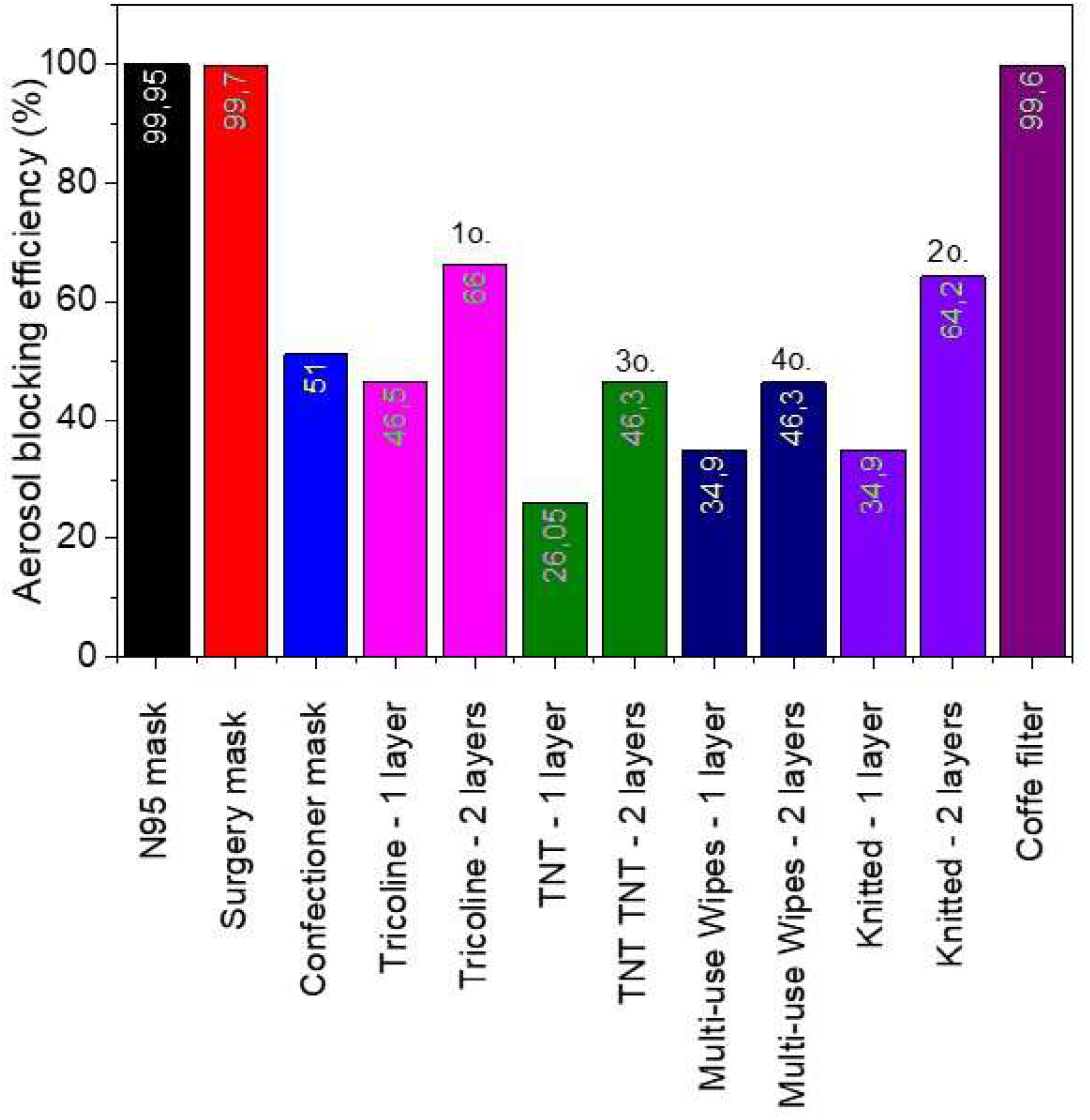
*The results were averaged from both techniques being associated with the right type of mask*.

In Figure 6, it can be seen that the N95 mask has a greater difficulty of air passage in relation to the other masks and fabrics, however the large effective area of the n95 mask reduces this effect and its discomfort. In general, masks and fabrics had small changes with increasing humidity, instead, the paper masks, due to their high capillarity and very small pores, blocked the aerosols but decreased the air passage. Water droplets being absorbed by the paper fibers covered the smaller pores due to its surface tension, showing a rapid variation in pressure as a function of exposure time to the nebulizer, thus representing a rapid increase in respiratory discomfort.

**Figure 6.**
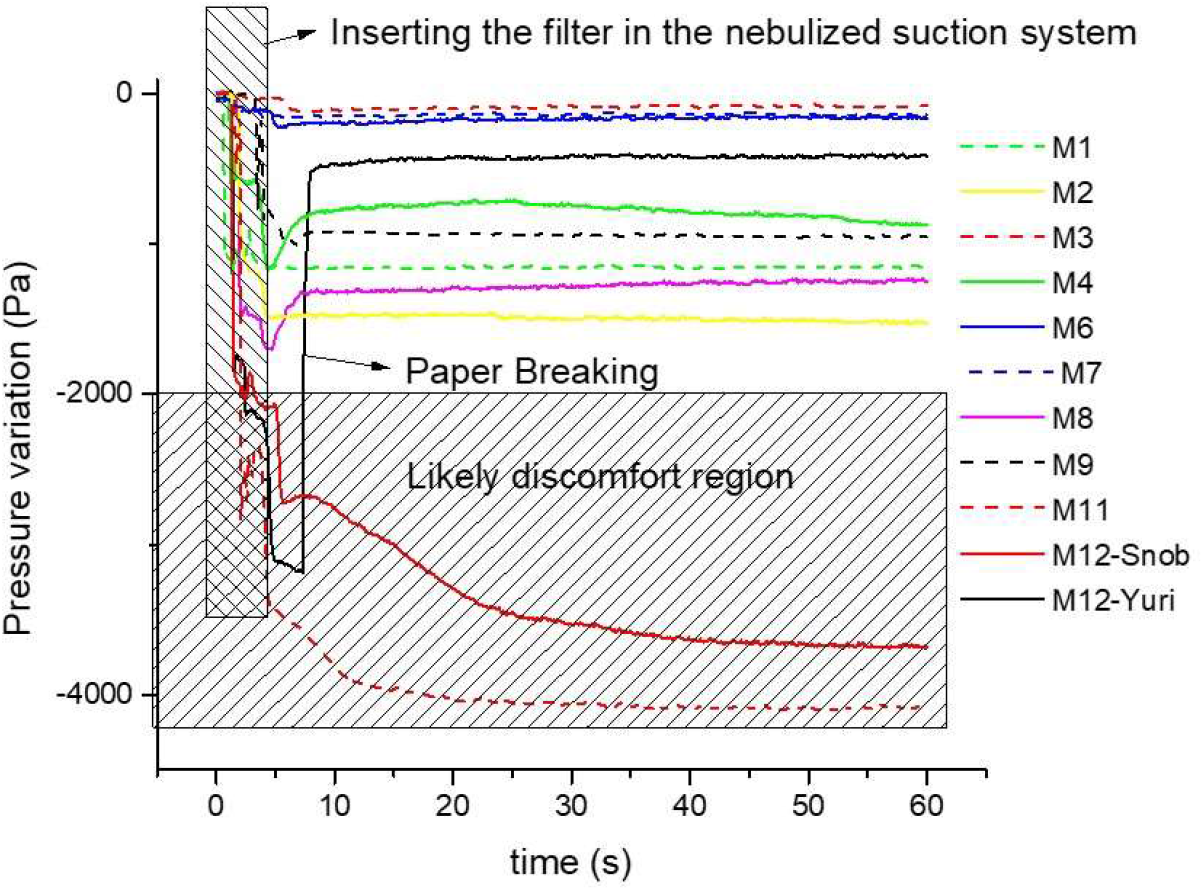
*Absolute pressure change as a function of time inside the aerosol line connecting the mask chamber*.

On carrying out pressure variation experiments, we also observed that flexible fabrics such as legging may deform itself very easily due to any pressure discontinuity, which could also increase the porosities and open the aerosol channels decreasing the effectiveness of the user protection. We also observed that one of the papers broke up with the increase in humidity, being able to increase the user risk. Therefore, all paper towels were discarded from use as layers in homemade masks. The paper filter used in coffee machines had higher air block, but also proved to be efficient for aerosol barrier, close to surgical masks. In the case of a filter #103, which has a large effective area, it reduces the effect of the pressure discontinuity, even so, it can create discomfort with the increase of humidity, consequently, it still can be used for a short period of time.

## CONCLUSIONS

The sort of masks used in this study played a central role in the characterization and validation directed to individual protection. Two different optical techniques and methodologies for characterizing respiratory masks has been presented. The quantitative scattering optical properties of aerosols passing through respiratory masks were obtained for a large batch fabrication, and the aerosol optical properties have demonstrated their suitability as a protection mean. SFDI was able to spatially localize the particles of aerosols. The results provide the groundwork for future studies on characterizing the passage of aerosols through respiratory masks with different preparations.

It is necessary to emphasize that the n95 and surgical masks are very stable with the humidity variation as can be seen in Figure 6, not changing the pressure difference as a function of time. As the n95 had 3 layers and the inner one is impermeable, it reduced the risk of particle transfer through the capillarities, defining this maks as the one with high protection and long use.

Note that the accommodation of the mask on the face is another important factor for increasing the efficiency of the masks, by avoiding the passage of aerosols through small cracks^14^. Finally, the making of homemade masks must be done according to the sequence informed in Figure 5, always giving preference to a greater number of layers, with woven fabrics with less porosity and with a higher percentage of cotton.

Note the individuals should preferably use the fabric masks, since the surgery masks should be available for health workers ^15^, furthermore the homemade fabric masks do not provide enough protection for the front group, i.e., the health workers ^16^

## Data Availability

The authors confirm that the data supporting the findings of this study are available within the article.

## ACKNOWLEDGMENTS

This work was supported by CNPq.

“This is a non-final version of an article to be published in a journal”.

## Notes

### Competing Interest Statement

The authors have declared no competing interest.

### Funding Statement

This work was supported by CNPq-Brazil.

